# Development of a health literacy prediction model: Using HLS-EU-Q16 questionnaire data from a population-based survey

**DOI:** 10.1101/2025.11.26.25341138

**Authors:** Suwon Hwang, Mankyu Choi

## Abstract

**Background:** Health literacy (HLIT) has been linked to both individual- and population-level outcomes; however, the large-scale measurement of HLIT presents practical challenges. This study aimed to develop a predictive model for HLIT using the 16-item European Health Literacy Survey Questionnaire (HLS-EU-Q16), a standardised survey instrument created by the HLS-EU consortium, to facilitate assessment and comparison of HLIT across countries.

**Methods:** Data were derived from the Korea Health Panel Survey, conducted between March and July 2021, involving a final sample selection of 9,510 participants. Variables were selected based on previously demonstrated theoretical and empirical relevance, and backward multiple linear regression was conducted to develop the prediction model on 70% of the data, which was later validated on the remaining 30%.

**Results:** The final model identified age, sex, education level, and annual household income as significant predictors of HLIT scores while maintaining model parsimony. The model explained approximately one-third of the variance in HLIT within the test set with a weighted R^2^ of 0.34 and showed a root-mean-square error of approximately 3.73 points in predicting the 0–16 HLIT score. Pearson’s correlation analysis further supported the validity of the model, showing a strong positive correlation between the observed and predicted HLIT scores (r = 0.63, p < 0.001).

**Conclusion:** The findings of our study provide a feasible model for predicting HLIT scores from population-based data, in this case, Korea Health Panel Survey data in combination with the HLS-EU-Q16, allowing for the effective measurement of HLIT, which would otherwise be time-consuming. This model has implications for use in cross-country comparisons of HLIT and in helping to inform public health policies and planning.

## BACKGROUND

Health literacy (HLIT) refers to an individual’s capacity to obtain, process, and understand health information to make appropriate health decisions. It encompasses a broad set of skills needed to access, comprehend, appraise, and apply information across health management, disease prevention, and health promotion domains [1]. However, ample evidence indicates that inadequate HLIT is common and linked to adverse outcomes. For example, nearly half of adults in developed countries have limited HLIT, which is associated with poorer health status and higher hospitalisation rates [2–6]. In Korea, nearly one in two adults has a limited level of HLIT, which is a concerning figure, given the nation’s high educational attainment and universal health coverage [7,8]. Low HLIT not only affects individual health behaviours and outcomes but can also exacerbate health disparities at the population level [9,10].

Despite its importance, the large-scale assessment of HLIT presents practical challenges. Standard instruments, such as the 16-item European Health Literacy Survey Questionnaire (HLS-EU-Q16) developed by the HLS-EU consortium to assess and compare HLIT across countries, provide a comprehensive measure of HLIT [5,10,11]. However, administering such questionnaires in every survey or clinical setting is time consuming. Population-wide HLIT assessments require substantial resources, and are infrequently conducted [5,7,13]. Consequently, policymakers and practitioners often use proxy indicators such as education level or income as rough surrogates for HLIT [14]. However, relying on a single proxy can misestimate an individual’s actual HLIT, as it overlooks the multifaceted nature of HLIT and the combined influence of multiple factors [5,15,16]. There is a need for predictive models that can integrate several demographic predictors to estimate HLIT more accurately without direct testing.

Previous attempts to develop such predictive models have been based primarily on data from the United States (U.S) and Europe, utilising either the U.S. version of HLIT assessments or the longer 47-item European Health Literacy Survey Questionnaire (HLS-EU-Q47) [17,18]. However, the literature in this area remains limited despite the substantial potential of prediction models to provide valuable estimates of HLIT at the population level [19]. Notably, South Korea has only recently begun carrying out comprehensive assessments of population HLIT, and to date, no study has attempted to create a predictive model specifically in relation to the shorter and more practical HLS-EU-Q16 instrument [7]. This study addresses this critical research gap by formulating and validating a predictive model of HLIT, using demographic and socioeconomic predictors based on scores from the HLS-EU-Q16, for comprehensive assessments, especially in Asian context such as Korea.

Recent studies have explored multivariate approaches for predicting HLIT. Studies have consistently shown that sociodemographic characteristics, such as older age, lower educational attainment, and lower income are associated with poorer HLIT [5,9,17,18]. For example, older age and lower educational levels can be linked to difficulties in understanding health information [20,21]. Combining such predictors in a model can improve estimation; a U.S. study demonstrated that a linear model with demographics accounted for approximately 30% of the variance in HLIT skills [17]. Similarly, in Europe, prediction models for HLIT have been developed and validated using census-like data, with education found to be a strong predictor, and showing that population HLIT levels can be satisfactorily estimated from a few key variables [18]. These efforts suggest that predictive modelling is a feasible strategy for gauging community HLIT and identifying at-risk groups without direct measurements.

Building on prior work, this study focused on the Korean context to develop a HLIT prediction model. Specifically, we used data from the Korea Health Panel Survey (KHPS) that incorporated the HLS-EU-Q16 instrument to measure HLIT at the national level [22].

## METHODS

### Aim

We aimed to construct a regression model using variables with previously demonstrated theoretical and empirical relevance and relationships with HLIT. The objectives of this study were two-fold: (1) to build a robust model that can predict an individual’s HLIT score based on basic demographic attributes, and (2) to validate the model’s performance and potential for application in estimating HLIT in population-based data. In doing so, we aimed to provide a practical tool for health authorities to identify communities with low HLIT and inform targeted health education and communication strategies.

### Data collection

This study utilised data from the KHPS, a nationally representative survey that examines factors influencing healthcare use, including socioeconomic characteristics, comorbidities, and health behaviours [22]. Participants in the KHPS were selected using the 2016 registered census as the sampling frame to reflect the changed population structure, and a two-stage clustered random sampling design to ensure representativeness. Individual sampling weights representing the inverse of the selection probability are present in the KHPS. This allowed us to derive national-level estimates and were included in our analysis. The survey was administered through computer-assisted personal interviews, and a wide range of information was obtained on health service utilisation, expenditure, and health status, as well as the sociodemographic characteristics of all participants. For this analysis, we used the most recent wave (2021), in which HLIT was assessed using the HLS-EU-Q16. Of the 11,057 responses, participants under 19 years of age were excluded, and those with valid responses to all 16 questions in the HLIT questionnaire were selected. The final sample size comprised 9,510 participants.

### Measures Dependent variable HLIT

HLIT was measured using the HLS-EU-Q16. This instrument covers three HLIT domains: health management (seven items), disease prevention (five items), and health promotion (four items). Each item asks respondents to rate how easy or difficult it is to find, understand, appraise, or apply certain types of health information (e.g. understanding instructions from a doctor or finding information about managing illness). Items are scored on a 4-point Likert scale (“very difficult”, “difficult”, “easy”, “very easy”), which are dichotomized for scoring such that “easy” and “very easy” responses receive 1 point and “difficult” or “very difficult” responses receive 0 points. The points are summed across the 16 items to produce a HLIT score ranging from 0 to 16, with higher scores indicating better-perceived HLIT. Following established conventions, scores are classified as 0–8 indicating “inadequate”, 9–12 indicating “marginal”, and 13–16 indicating “adequate” HLIT [23]. The HLS-EU-Q16 has been validated in multiple languages and contexts; notably, a Korean version of this instrument has been developed with a forward-backward translation process and has demonstrated good internal consistency (Cronbach’s α = 0.86) in the Korean population [12]. The validated Korean HLS-EU-Q16 was implemented in the 2021 KHPS to gauge respondents’ HLIT and provide an outcome measure for our model.

### Independent Variables (Predictors)

Based on the literature and available data, we included age, sex, marital status, residential area, economic activity, level of education, and annual household income as predictors in the initial model [3,4,15,21]. Age was measured in years as a continuous variable and centred at the mean (approximately 50 years) for regression analyses to improve the interpretability of the intercept. Marital status was categorised as either married or single (which included being single, divorced, separated, or widowed). Residential areas were classified as urban or rural, and economic activity was categorised as either currently employed or not. Education was categorised as: (1) middle school or lower, (2) high school, or (3) college degree or higher. For regression modelling, we treated education as an ordinal indicator of educational attainment, with higher values corresponding to higher levels of education. Annual household income in Korean won (KRW) was reported in the survey. Given the right-skewed income distribution, we applied a logarithmic transformation (natural log of annual income) to reduce skewness and linearise the relationship with HLIT. In addition, income quartiles (Q1=lowest 25% through Q4=highest 25%) were categorised for descriptive analysis. No other demographic variables (such as ethnicity or language) were available in the KHPS as the Korean population is largely ethnically homogeneous.

### Data Analysis and Model Specification

Data were analysed using R 4.4.2 software. We specified a multiple linear regression model with the HLIT score as the dependent variable and the aforementioned predictors as the independent variables. The dataset was randomly split into two parts: one containing 70% of the data (N = 6,657) to formulate our prediction model, and the other 30% (N= 2,853) to test our prediction model. A backward multiple linear regression was carried out to construct our prediction model, where variables that were not significantly associated (and excluding the least significant variable after for model parsimony) with HLIT were excluded step by step. The model is formed as follows:

Equation 1. The HLIT prediction model

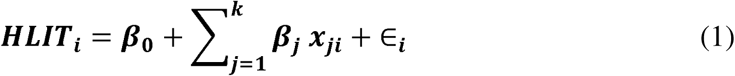

where HLIT_i_ is the observed HLIT score for the ith individual, p_O_ is the intercept term, x_ji_ represents the value of the jth predictor for individual;, p_j_ refers to the regression coefficients associated with each predictor variable included in the model, and E_i_ is the error term for individual *i*.

We reported the coefficient of determination (R^2^) as a measure of explained variance in the training set, weighted R^2^ to adjust for the survey design and model performance in the test set, and the root mean square error (RMSE) as a measure of prediction error in the same units as the HLIT score. The statistical significance of each coefficient was evaluated at a 95% confidence level (α = 0.05). All analyses were conducted using the individual survey weights present in the KHPS data.

### Validation of the Prediction Model

A cross-validation approach was used to validate the prediction model. The model was fitted to the training set, and the resulting equation was used to predict the HLIT scores for individuals in the test set. We then calculated the R^2^, weighted R^2^, and RMSE of the test set predictions to assess how well the model generalised to the new data. In addition, we performed a Pearson’s correlation test to examine the association between the observed and predicted HLIT scores. We conducted standard regression diagnostics to ensure the validity of our model and computed the variance inflation factor (VIF) for each predictor to detect multicollinearity between the independent variables.

## RESULTS

Table 1 presents the descriptive characteristics of the sample used to formulate the prediction model (N = 6,657). The proportion of females was 53.0%, and the mean age was approximately 50 years. Most individuals were married, lived in urban regions, were economically active, had a college degree or higher, and were in the highest (Q4) income quartile.

**Table 1.**
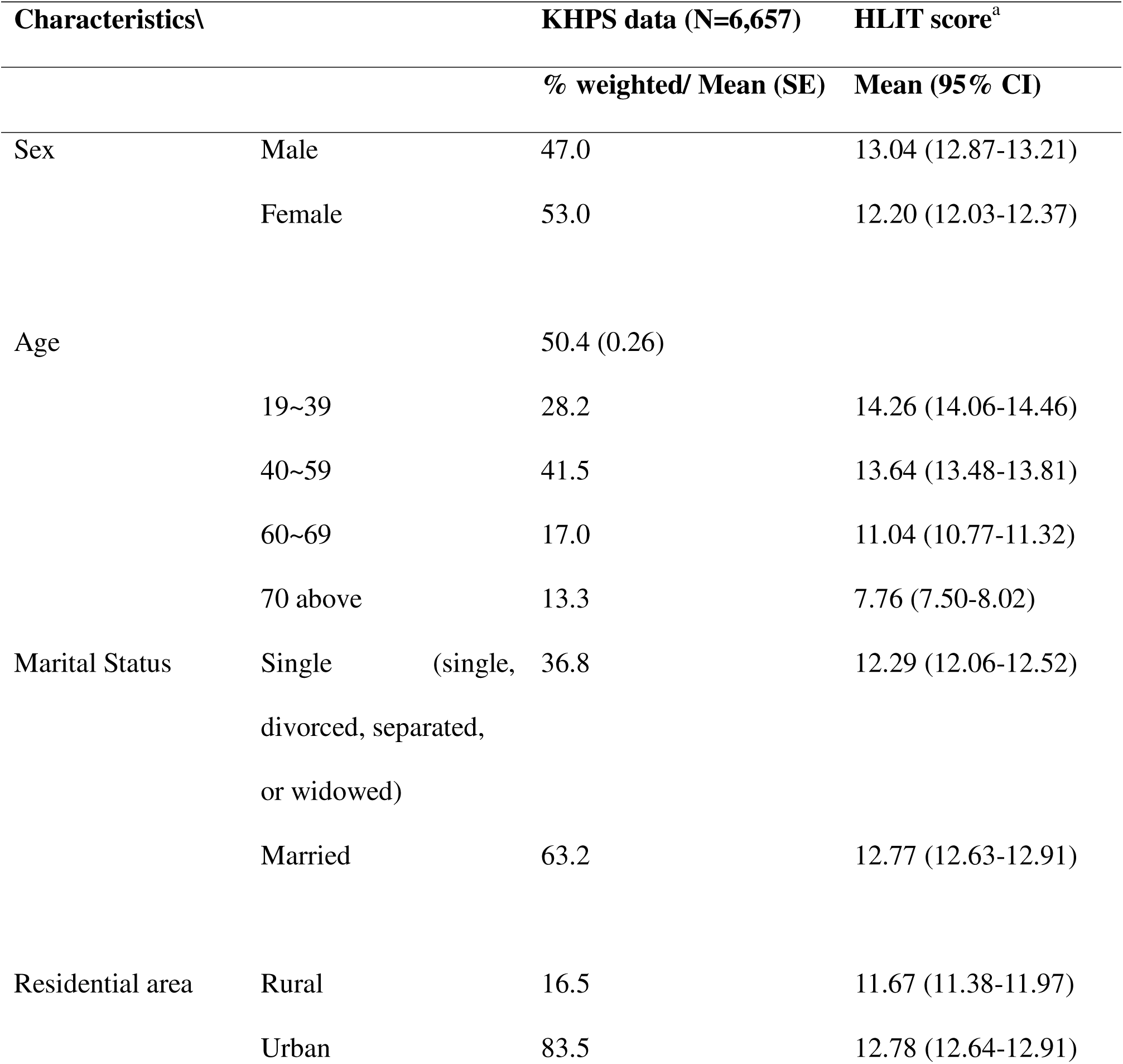
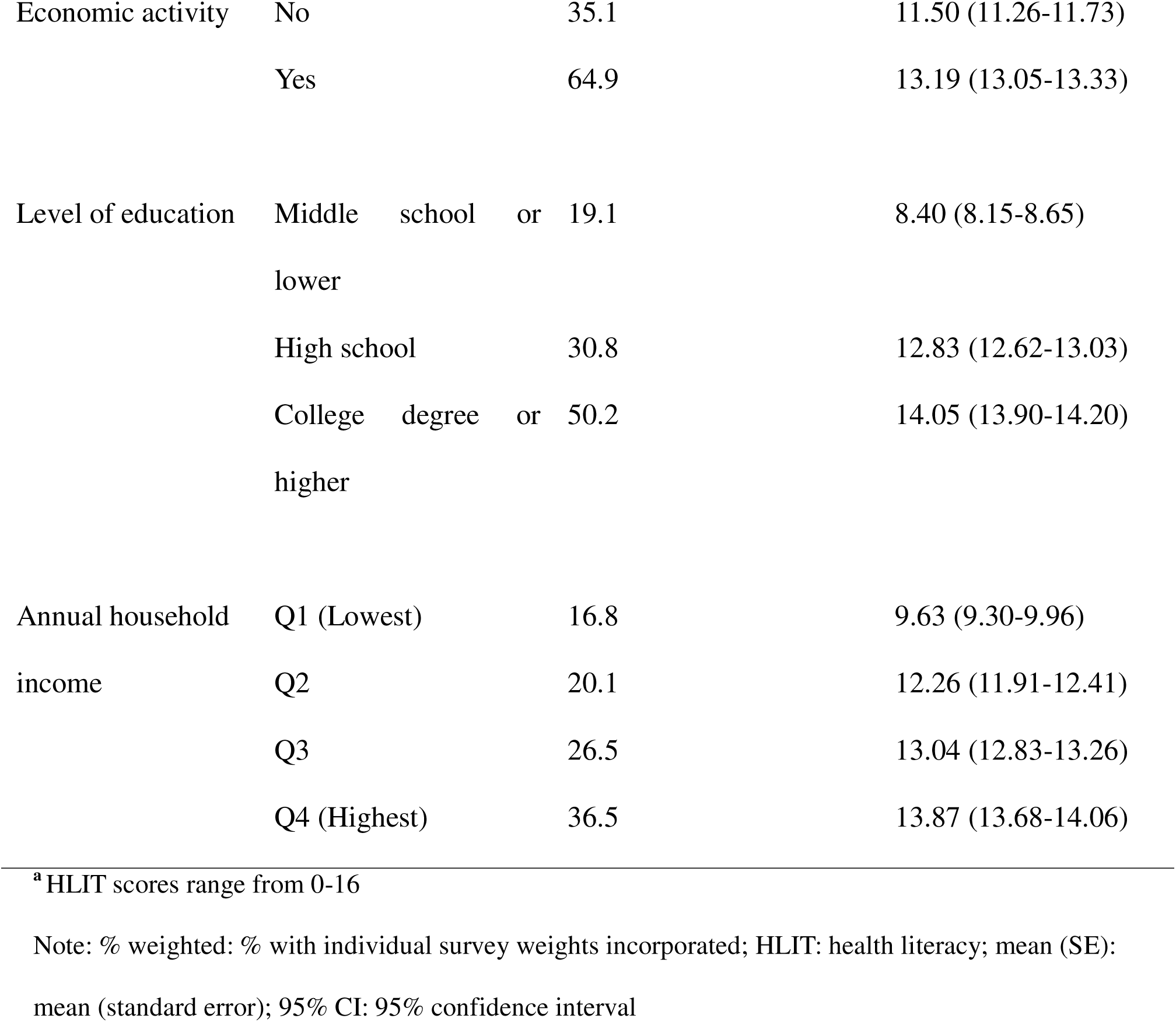
Sample characteristics and distribution of mean HLIT scores based on 70% of the KHPS data.

| Characteristics\ |  | KHPS data (N=6,657) | HLIT score<sup>a</sup> |
| --- | --- | --- | --- |
|  |  | % weighted/ Mean (SE) | Mean (95% CI) |
| Sex | Male | 47.0 | 13.04 (12.87-13.21) |
|  | Female | 53.0 | 12.20 (12.03-12.37) |
| Age |  | 50.4 (0.26) |  |
|  | 19~39 | 28.2 | 14.26 (14.06-14.46) |
|  | 40~59 | 41.5 | 13.64 (13.48-13.81) |
|  | 60~69 | 17.0 | 11.04 (10.77-11.32) |
|  | 70 above | 13.3 | 7.76 (7.50-8.02) |
| Marital Status | Single (single, divorced, separated, or widowed) | 36.8 | 12.29 (12.06-12.52) |
|  | Married | 63.2 | 12.77 (12.63-12.91) |
| Residential area | Rural | 16.5 | 11.67 (11.38-11.97) |
|  | Urban | 83.5 | 12.78 (12.64-12.91) |
| Economic activity | No | 35.1 | 11.50 (11.26-11.73) |
|  | Yes | 64.9 | 13.19 (13.05-13.33) |
| Level of education | Middle school or lower | 19.1 | 8.40 (8.15-8.65) |
|  | High school | 30.8 | 12.83 (12.62-13.03) |
|  | College degree or higher | 50.2 | 14.05 (13.90-14.20) |
| Annual household income | Q1 (Lowest) | 16.8 | 9.63 (9.30-9.96) |
|  | Q2 | 20.1 | 12.26 (11.91-12.41) |
|  | Q3 | 26.5 | 13.04 (12.83-13.26) |
|  | Q4 (Highest) | 36.5 | 13.87 (13.68-14.06) |

<sup>a</sup> HLIT scores range from 0-16
Note: % weighted: % with individual survey weights incorporated; HLIT: health literacy; mean (SE): mean (standard error); 95% CI: 95% confidence interval

Table 2 presents the initial and final prediction models for HLIT fitted to the training data. Residential area was insignificant in our initial model and was thus excluded. In addition, marital status and economic activity were excluded from the final model because of their minimal significance and to enhance model parsimony. Sex, age, educational level, and annual household income were the remaining significant predictors of HLIT in the final model. Being female, older, and having lower education levels were associated with lower HLIT scores, whereas a higher annual household income was associated with a higher HLIT score. The final model achieved an R^2^ of 0.31 on the training set, indicating that 31% of the variance in HLIT could be explained by the selected predictors. Furthermore, the adjusted R^2^ value (0.31) accounted for the number of predictors, suggesting a well-fitted model. For the test data, the weighted R^2^ was 0.34, indicating that the model generalised well and explained 34% of the variance in the HLIT scores among individuals. The RMSE was 3.73, suggesting a moderate level of prediction accuracy. All VIFs were below 2.2, indicating that, while the independent variables correlated to some extent, they did not pose multicollinearity concerns in the model.

**Table 2.**
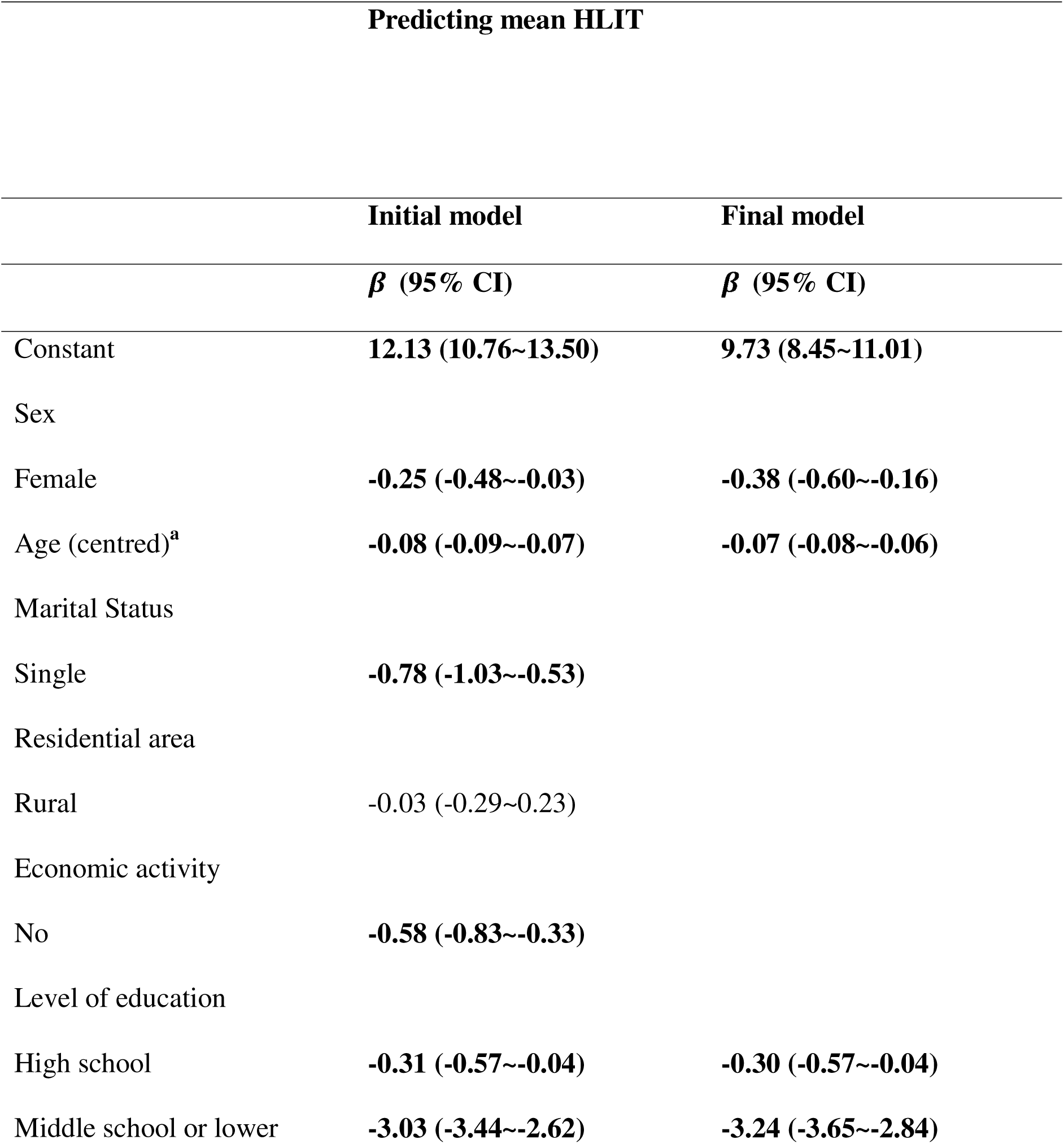
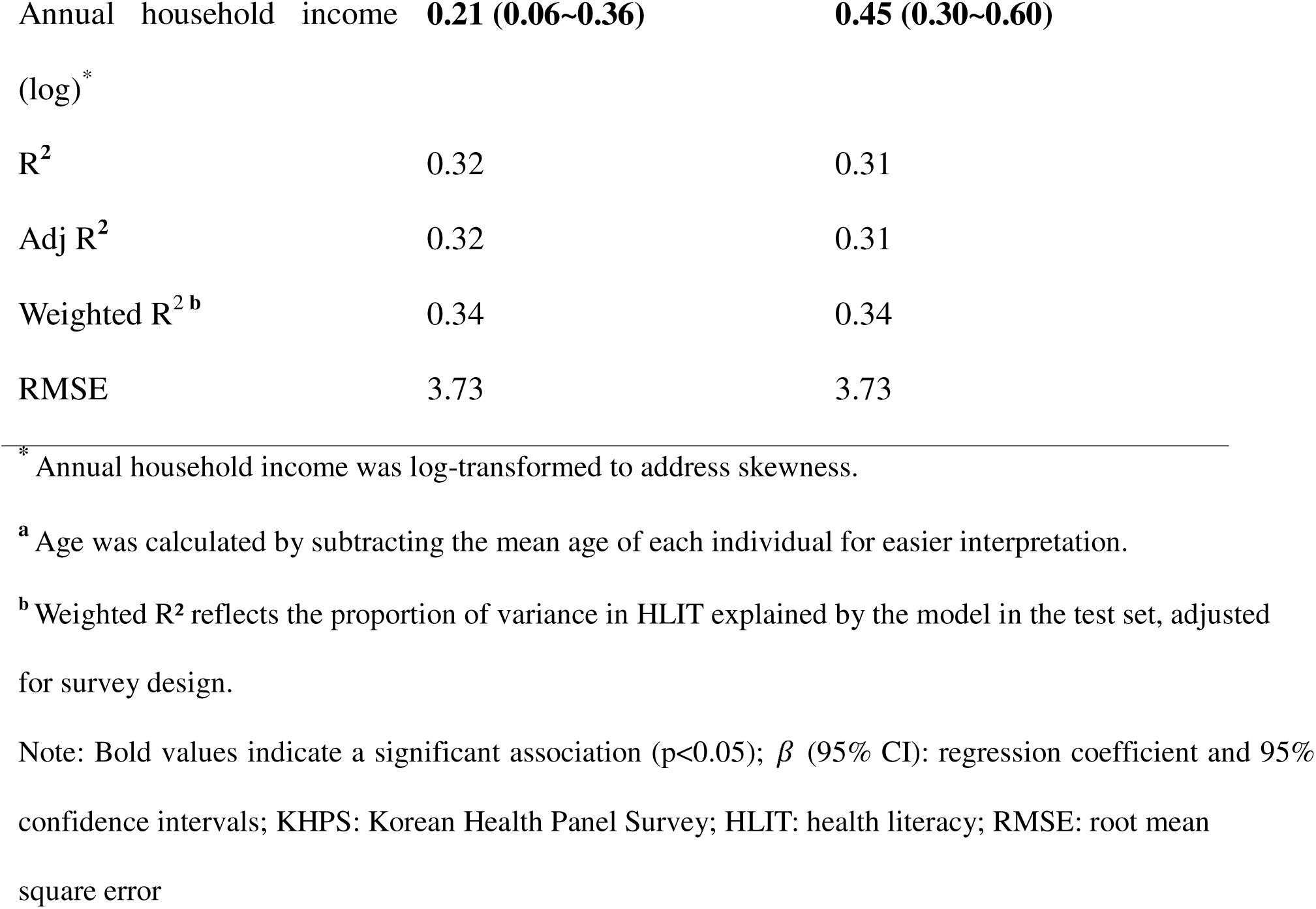
The HLIT prediction model based on 70% of the KHPS dataset (N=6,657) and validation based on test data (N=2,853)

Table 3 presents the observed and predicted mean HLIT scores per decile, demonstrating that the HLIT can accurately predict subpopulations (see Figure 1). Pearson’s correlation analysis further supported the validity of the model, showing a strong positive correlation between the observed and predicted mean HLIT scores (r = 0.63, p < 0.001), confirming that the model provided meaningful estimates of HLIT levels.

**Figure 1:**
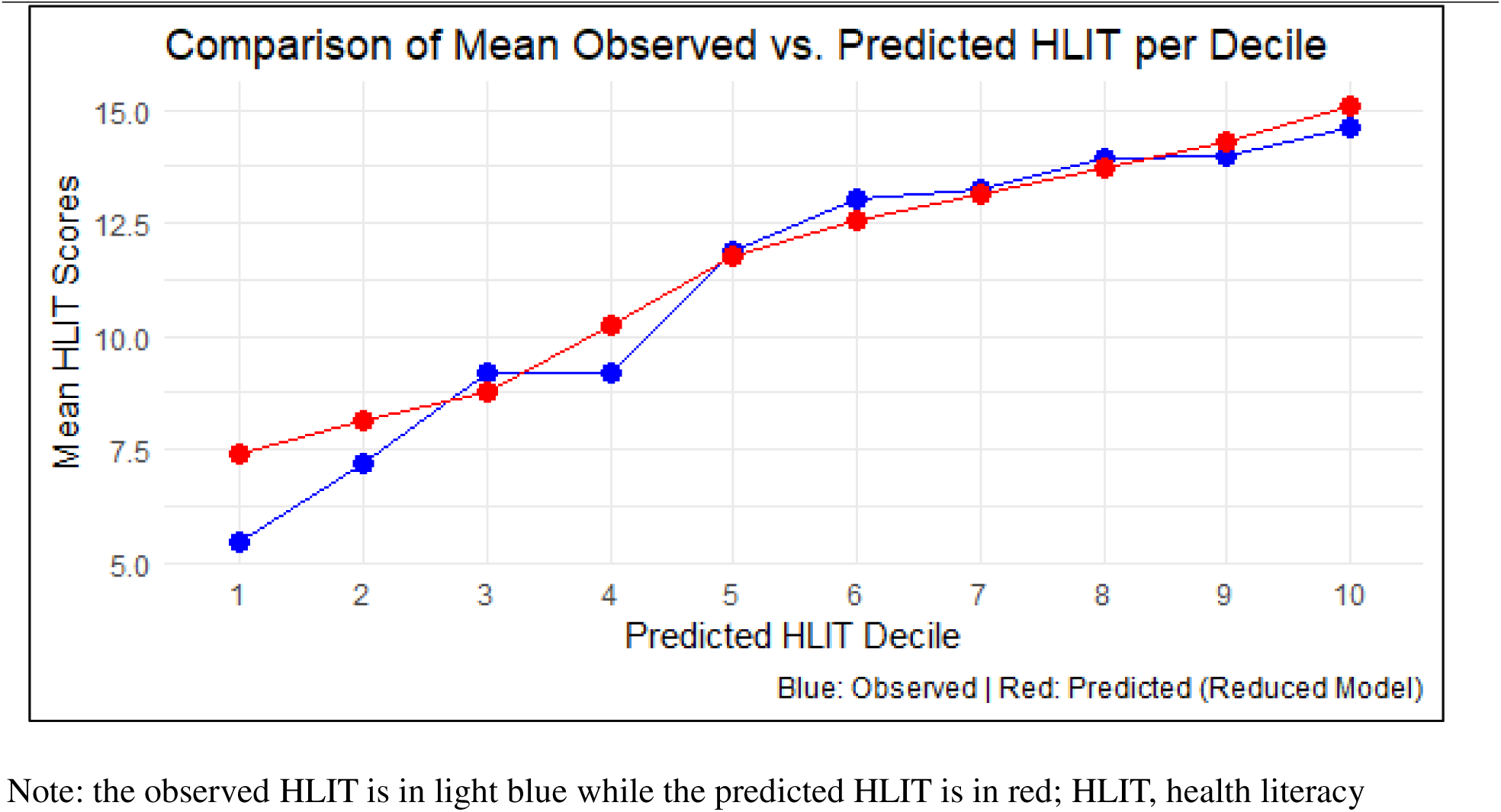
Observed and predicted mean health literacy score per decile

**Table 3.** Observed and predicted mean HLIT scores overall and per decile based on the KHPS data.

| Per decile | Observed mean HLIT score | Predicted mean HLIT score |
| --- | --- | --- |
|  | Mean (SE) | Mean (SE) |
| **Total** | 12.59 (0.09)<sup>a</sup> | 12.65 (0.05)<sup>a</sup> |
| 1 | 5.43 (0.21) | 7.40 (0.02) |
| 2 | 7.19 (0.24) | 8.16 (0.01) |
| 3 | 9.17 (0.24) | 8.78 (0.01) |
| 4 | 9.18 (0.27) | 10.3 (0.04) |
| 5 | 11.9 (0.25) | 11.8 (0.02) |
| 6 | 13.0 (0.22) | 12.5 (0.01) |
| 7 | 13.2 (0.21) | 13.1 (0.01) |
| 8 | 13.9 (0.19) | 13.7 (0.01) |
| 9 | 14.0 (0.18) | 14.3 (0.01) |
| 10 | 14.6 (0.14) | 15.1 (0.02) |

<sup>a</sup> $r = 0.630$ ( $p < 0.001$ )
Note: Mean (SE): mean and standard error; KHPS: Korean Health Panel Survey; HLIT: health literacy

## DISCUSSION

This study aimed to develop a predictive model for HLIT scores based on nationally representative survey data using the HLS-EU-Q16. Our findings indicated that age, sex, educational level, and annual household income were significant predictors of HLIT while maintaining model parsimony. The findings of this study concerning this model offer a means for predicting HLIT scores from population-based data, allowing for the measurement of HLIT, which would otherwise be time-consuming.

Our findings align with those of previous studies, reaffirming that key sociodemographic variables are strongly associated with HLIT. Aging has consistently been linked to lower HLIT because cognitive and functional impairments can hinder the ability to access, process, and utilise health information and services effectively [20]. Similarly, higher levels of education and income serve as crucial enablers, facilitating both comprehension of health-related information and access to quality healthcare resources [21]. The well-established relationship between HLIT and these factors remained evident in our predictive model, demonstrating its robustness across different contexts, including the Korean population, irrespective of the specific HLIT assessment instrument employed.

Overall, our model demonstrated moderate ability to predict mean HLIT scores, underscoring the broader potential of predictive models for estimating HLIT at a large-scale population level using national census datasets. Such models provide practical alternatives to direct assessments, enabling more efficient resource allocation and policy planning [14]. For example, Rasu et al. applied Martin’s model to a nationally representative dataset to assess the impact of HLIT on healthcare utilisation and expenditure [17,19]. Although our prediction model could not be validated in other countries due to the limited availability of population-wide HLS-EU-Q16 surveys, our findings align closely with those of Martin and Heide’s HLIT predictive model, suggesting its broader applicability across different contexts [13,17,18].

Although prior efforts to develop HLIT prediction models have been made in the U.S. and Europe, to the best of our knowledge, this study is the first to construct such a model in an Asian context. Notably, a study validating the extended HLS-EU-Q47 across six Asian countries also found strong associations between HLIT and factors such as educational attainment and social status [13]. Moreover, research has demonstrated that the HLS-EU-Q47 could capture the relationship between HLIT and its determinants in a manner consistent with earlier findings from Western contexts [3,5]. These parallels suggest that the underlying predictors of HLIT may be comparable across diverse cultural settings, which is further supported by the prediction model in the present study.

This study has several limitations. First, the HLS-EU-Q16 assesses self-reported HLIT, which may not fully capture an individual’s actual HLIT level owing to potential biases in self-assessment. Nevertheless, it remains a widely accepted and practical tool for measuring perceived HLIT across different populations. Second, the predictive model developed in this study could not be externally validated using other large-scale population datasets, either within Korea or internationally because of the limited availability of comparable surveys. This restricts the generalisability of the model and highlights the need for further validation across diverse populations and settings.

## CONCLUSION

The findings of our study show that HLIT levels can be predicted from population-based data, helping to inform public health policies and planning. By identifying key sociodemographic predictors such as age, sex, education, and income, the model offers a practical solution in situations where direct HLIT measurement is not possible due to time or resource limitations. This approach provides health authorities with a useful tool to assess population-level HLIT, identify at-risk groups, and design targeted interventions. Integrating such models into existing surveys or census data can improve the reach and fairness of health strategies. Although the model showed moderate predictive strength, its consistency with international findings suggests that it can be adapted for wider use. Future studies should validate the model in different populations and explore ways to improve its accuracy by including additional factors. Overall, this study offers a simple and scalable method for estimating HLIT, supporting better targeted public health decision-making.

## LIST OF ABBREVIATIONS

HLIT: Health literacy
HLS-EU-Q16: European Health Literacy Survey Questionnaire

## Declarations

### Ethics approval and consent to participate

This study adhered to the Declaration of Helsinki and the Institutional Review Board of Korea University approved the study protocol, granting an IRB exemption and waiving the requirement for informed consent (reference No. KU_IRB-2025-0153-01).

### Consent for publication

Not applicable.

### Competing interests

The authors have declared that no competing interests exist.

## Funding

The authors received no specific funding for this work

## Authors’ contributions

Conceptualization: S.H; Data curation: S.H; Formal analysis: S.H; Writing – original draft: S.H; Writing – reviewing & editing: S.H, M.K; supervision: M.K

All authors read and approved the final version of the manuscript.

## Data Availability

The data is owned by a third party and the authors had no special access privileges others would not have. A request for the data used for this study can be made on https://www.khp.re.kr:444/eng/data/data.do. Inquiries regarding data acquisition should be sent to the Korea Institute for Health and Social Affairs.

## Acknowledgements

Not applicable.

## Clinical trial number

Not applicable.

## Notes

### Competing Interest Statement

The authors have declared no competing interest.

### Funding Statement

This study did not receive any funding

### Author Declarations

This study adhered to the Declaration of Helsinki and the Institutional Review Board of Korea University approved the study protocol, granting an IRB exemption and waiving the requirement for informed consent (reference No. KU_IRB-2025-0153-01). The data is owned by a third party and the authors had no special access privileges others would not have. A request for the data used for this study can be made on https://www.khp.re.kr:444/eng/data/data.do. Inquiries regarding data acquisition should be sent to the Korea Institute for Health and Social Affairs.

